# Artificial Intelligence-Large Language Models (AI-LLMs) for Reliable and Accurate Cardiotocography (CTG) Interpretation in Obstetric Practice

**DOI:** 10.1101/2024.11.13.24317298

**Authors:** Khanisyah Erza Gumilar, Manggala Pasca Wardhana, Muhammad Ilham Aldika Akbar, Agung Sunarko Putra, Dharma Putra Perjuangan Banjarnahor, Ryan Saktika Mulyana, Ita Fatati, Zih-Ying Yu, Yu-Cheng Hsu, Erry Gumilar Dachlan, Chien-Hsing Lu, Li-Na Liao, Ming Tan

## Abstract

**BACKGROUND:** Accurate interpretation of Cardiotocography (CTG) is a critical tool for monitoring fetal well-being during pregnancy and labor, providing crucial insights into fetal heart rate and uterine contractions. Advanced artificial intelligence (AI) tools such as AI-Large Language Models (AI-LLMs) may enhance the accuracy of CTG interpretation, leading to better clinical outcomes. However, this potential has not yet been examined and reported yet.

**OBJECTIVE:** This study aimed to evaluate the performance of three AI-LLMs (ChatGPT-4o, Gemini Advance, and Copilot) in interpreting CTG images, comparing their performance to junior and senior human doctors, and assessing their reliability in assisting clinical decisions. STUDY DESIGN: Seven CTG images were evaluated by three AI-LLMs, five senior doctors (SHD), and five junior doctors (JHD) and rated by five maternal-fetal medicine (MFM) experts (raters) using five parameters (relevance, clarity, depth, focus, and coherence). The raters were blinded to the source of interpretations, and a Likert scale was used to score the performance of each system. Statistical analysis assessed the homogeneity of expert ratings and the comparative performance of AI-LLMs and doctors.

**RESULTS:** ChatGPT-4o outperformed the other AI models with a score of 77.86, much higher than Gemini Advance (57.14) and Copilot (47.29), as well as the junior doctors (JHD; 61.57). CG4o’s performance (77.86) was only slightly below that of the senior doctor (SHD; 80.43), with no statistically significant differences between CG4o and SHD (p>0.05). Meanwhile, CG4o had the greatest score in the “depth” category, while the other four parameters were only marginally behind SHD.

**CONCLUSION:** CG4o demonstrated outstanding performance in CTG interpretation, surpassing junior doctors and other AI-LLMs, while senior doctors remain superior in all groups. AI-LLMs, particularly CG4o, showed promising potential as valuable tools in clinical practice to assist obstetricians, enhance diagnostic accuracy, and improve patient care.

## Introduction

Cardiotocography (CTG) is a vital monitoring method in modern obstetrics to assess the condition of the fetus during labor[1]. CTG monitors both the fetal heart rate and uterine contractions simultaneously, providing crucial information about fetal well-being. CTG serves as a valuable monitoring tool during pregnancy and labor, as it can detect fetal heart rate deceleration[2], hypoxia[3], and excessive contractions[4], all of which impact mother baby and mother safety. The presence of CTG can reduce labor complications, morbidity, and infant mortality[5]. Although its usage has become standard, interpreting CTG results often requires specialized training, experience, and expertise. A mistaken interpretation or delayed response can significantly impact both the mother and the fetus. Therefore, there is a pressing need for technology to enhance the accuracy and efficiency of CTG interpretation. The development of Artificial Intelligence-Large Language Models (AI-LLM) in recent years has opened new possibilities in various fields, including medicine. AI-LLMs can quickly and accurately analyze large volumes of data, learning from patterns and trends that humans may not easily detect[6]. In the context of CTG, AI-LLMs can provide rapid and accurate initial analyses, assisting medical professionals in making timely and data-driven decisions. The use of AI-LLMs in CTG interpretation offers several key advantages. These models are trained on large datasets of historical CTG data, enabling them to accurately identify patterns associated with specific risks. They are accessible anytime and anywhere, providing consistent support without fatigue or biases that could affect human interpretation. AI-LLMs also can learn and improve as more data becomes available continuously[7]. With each new interaction and interpretation, these models refine their algorithms, becoming more reliable and effective over time. Furthermore, integrating AI-LLMs into healthcare systems can reduce the workload of medical staff, allowing them to focus on other aspects of patient care that require more attention.

In the increasingly digital world, implementing AI-LLMs in CTG interpretation can be a significant innovation in obstetrics and childbirth practices, by providing fast, accurate, and consistent analysis, thus improving the safety of mothers and babies during labor. This research aims to assess the accuracy of interpretations of CTG images produced by different AI-LLMs for analyzing diverse patient conditions and determine whether the AI-LLMs are reliable in assisting doctors’ work. By combining medical expertise with advanced technology, our goal is to develop improved and more efficient solutions to ensure safe and optimal childbirth processes.

## Materials and Methods

### Materials

We used three AI-based chatbots in this study: ChatGPT-4o (https://chatgpt.com/), Gemini Advance (https://gemini.google.com/app), and Copilot (https://www.bing.com/). The CTG images used in this study were from the Department of Obstetrics and Gynecology, Universitas Airlangga Hospital, Surabaya, Indonesia (Ethical Approval from Universitas Airlangga Hospital no.156/KEP/2024; protocol number UA-02-24216). CTG-1 (Fig. 1A) was taken from a 37-week pregnant patient with preeclampsia and gestational diabetes undergoing induction with misoprostol. CTG-2 (Fig. 1B) is from a 38-week pregnant patient with a history of premature rupture of membranes (PROM). CTG-3 (Fig. 1C) was obtained from a 37-week and 5-day pregnant patient in active labor, where decelerations were observed. CTG-4 (Fig. 1D) is the CTG of a 39-week pregnant patient who underwent intrauterine resuscitation after a previous CTG indicated Category-2 or suspicious NST. CTG-5 (Fig. 1E) was taken from a 41-week post-date pregnant patient with PROM undergoing induction. CTG-6 (Fig. 1F) represents the CTG of a 37-week pregnant patient who presented with a 3-day history of fever. This patient was diagnosed with a severe respiratory tract infection. CTG-7 (Fig. 1G) is from a 41-week post-date pregnant patient undergoing induction with misoprostol.

**Figure 1.**
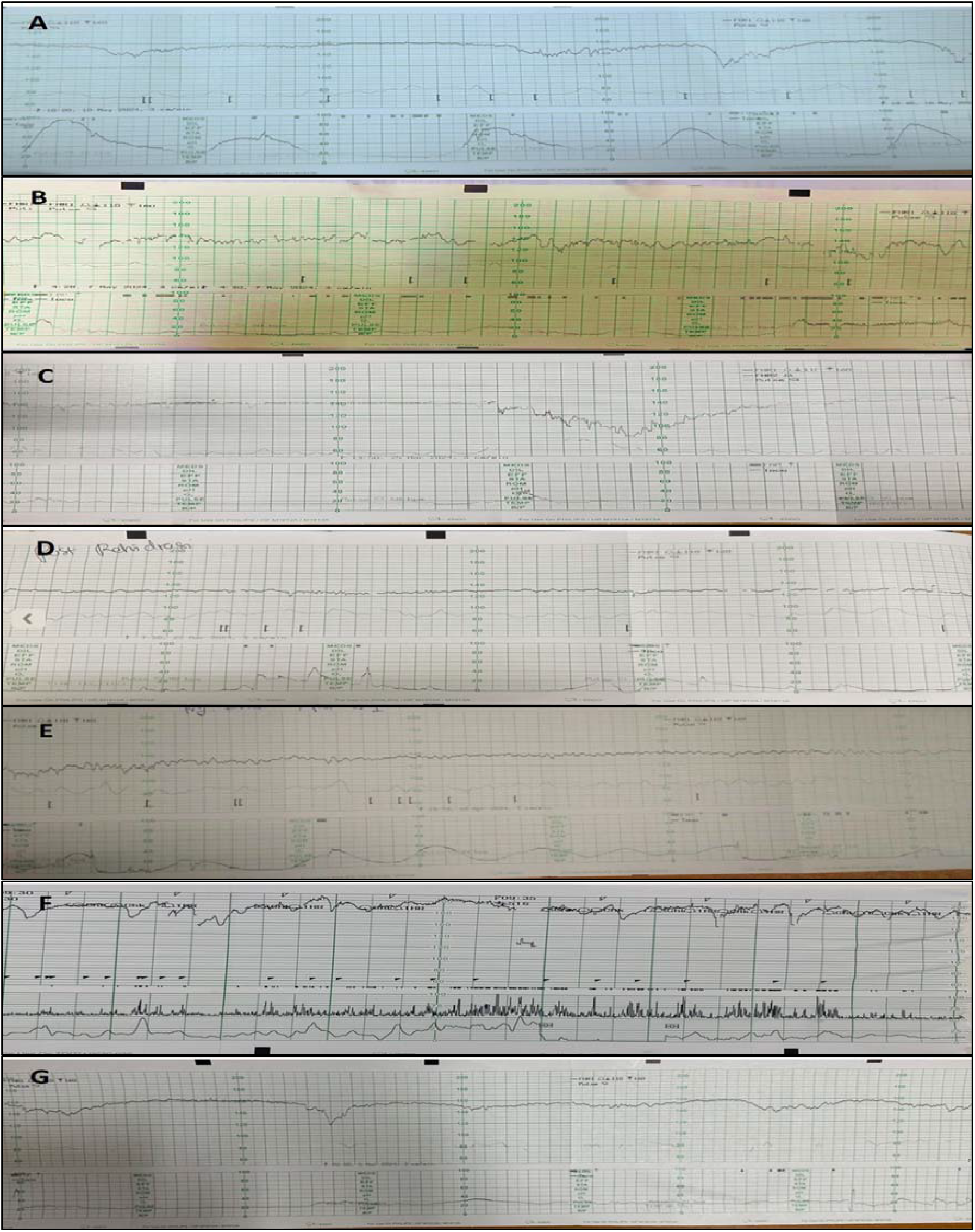
The CTG images used in this study. **A**. A 37-week pregnancy with preeclampsia and gestational diabetes undergoing misoprostol induction shows regular uterine contractions; **B**. A 38-week pregnancy with PROM without accompanying contractions; **C**. First stage of labor in the active phase experiencing secondary arrest and decelerations; **D. CTG condi**tion after intrauterine resuscitation shows low variability; **E**. Post-date pregnancy with PROM undergoing induction. The induction was eventually stopped due to meconium-stained amniotic fluid; **F**. A pregnant patient with severe symptomatic pneumonia and fever for 3 days before arriving at the hospital; **G**. Misoprostol induction in a post-date pregnancy at 41 weeks. After one series of administration,

We recruited 10 obstetrics doctors as participants for comparative assessment. Five physicians with more than 10 years of experience in obstetrics were categorized as senior human doctors (SHD), while the remaining five physicians with less than 5 years of obstetrics experience were grouped as junior human doctors (JHD). Additionally, we enlisted five maternal-fetal medicine (MFM) specialists to evaluate all the interpretations derived from the three AI-LLMs and the two groups of doctors.

### Study Design

We evaluated the performance of three LLMs (known as chatbots): ChatGPT-4o (referred to as CG4o), Gemini Advance (referred to as GemAdv), and Copilot, when presented with CTG images in labor cases. Both groups of doctors (SHD and JHD) were also tested with the same questions as those given to the AI-LLMs. Seven CTG images from different patients were presented to each chatbot, along with a specific prompt, and the interpretations were recorded (Fig. 2) (Supp.1). These interpretations were then reviewed by a team of 5 MFM (raters). To ensure impartiality, the responses from the chatbots were coded and randomized before being blindly assessed by the raters. The raters evaluated the responses without knowing which chatbot generated each result. To analyze the output of the chatbots, we used 5 parameters, including “relevance”, “clarity”, “depth”, “focus”, and “coherence” (Table 1) [8-12] with a 5-point Likert scale (Table 2) [13, 14].

**Table 1.** Parameters of assessment.

| Definition & Parameters |  |
| --- | --- |
| **Relevance:** | The response is closely related or appropriate to the issue |
| **Clarity:** | Clear, easy to understand, free from ambiguity |
| **Depth:** | The answer provides detailed and specific information, not just a general or superficial answer |
| **Focus:** | Contains the main points or keywords expected |
| **Coherence:** | All parts of the answer work together in a logical and structured way, with no conflicting parts |

**Table 2:**
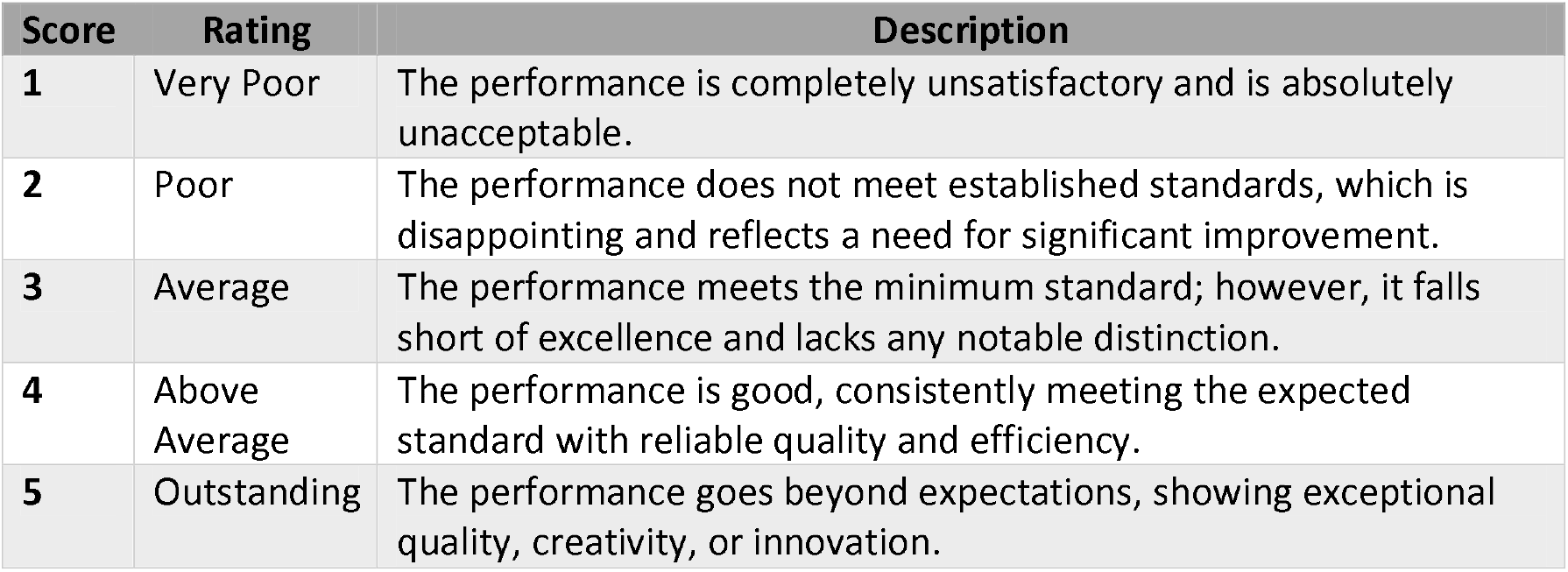
Likert scale rating and descriptions.

| Score | Rating | Description |
| --- | --- | --- |
| 1 | Very Poor | The performance is completely unsatisfactory and is absolutely unacceptable. |
| 2 | Poor | The performance does not meet established standards, which is disappointing and reflects a need for significant improvement. |
| 3 | Average | The performance meets the minimum standard; however, it falls short of excellence and lacks any notable distinction. |
| 4 | Above Average | The performance is good, consistently meeting the expected standard with reliable quality and efficiency. |
| 5 | Outstanding | The performance goes beyond expectations, showing exceptional quality, creativity, or innovation. |

**Figure 2.**
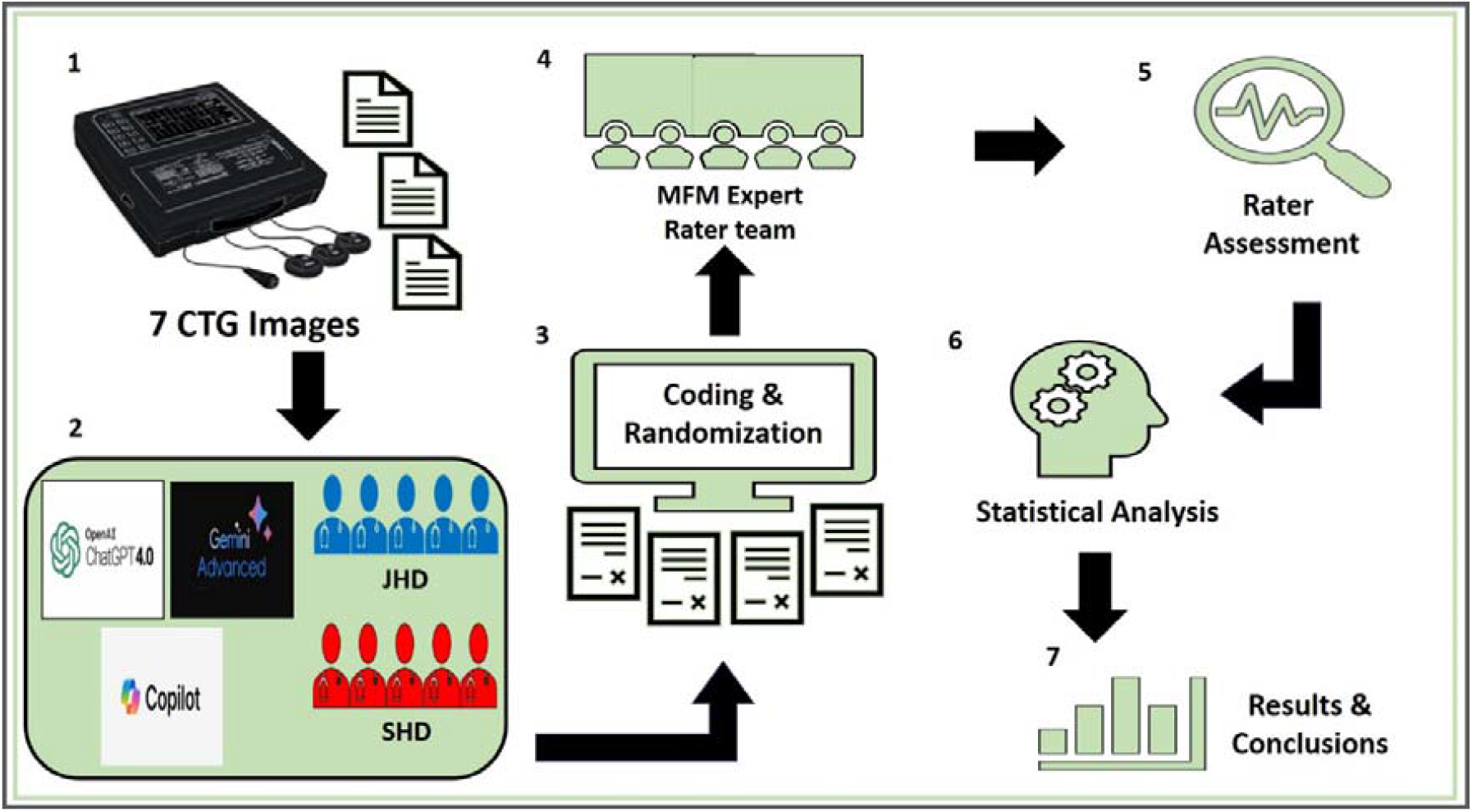
Study algorithm. Seven CTG images from labor cases (1) were tested on three groups (AI-LLM, JHD, and SHD) (2). The interpretations from these three groups were assigned special codes and randomized (3) for subsequent evaluation by MFM experts (4). The expert team assessed the CTG interpretations from various entities using agreed-upon parameters (5). The study results were managed through statistical analysis for evaluation and comparison (6), which were then presented in the form of figures and conclusion (7).

### Statistical analysis

This study aimed to evaluate the performance of three chatbots and human doctors in interpreting seven CTG images. Five raters were engaged to evaluate all responses generated by the three chatbots and human doctors using five parameters: relevance, clarity, depth, focus, and coherence. A 5-point Likert scale was employed for scoring, with a score of 5 indicating superior performance. Subsequently, the scores were linearly converted to a 0– 100 scale, following prior studies and recommendations[15]. To ascertain the extent of inter-rater consistency (homogeneity) among the five raters, the Pearson and Spearman correlation coefficients were employed as a means of quantifying the results. Furthermore, a one-way ANOVA with Scheffe’s post hoc analysis was utilized to investigate the differences between the mean scores of the raters. A multiple linear regression model was constructed to evaluate the performance of both chatbots and human doctors. This model accounts for the potential influence of subjectivity among raters and the varying degrees of complexity observed among image sets. All statistical analyses were conducted using the SAS software (Version 9.4, SAS Institute, Cary, NC, USA), with a significance level set at 0.05.

## Results

### Raters’ scores showed a moderate to high homogeneity

To assess the consistency of the raters’ scores for the CTG interpretation responses, we utilized a heat map to illustrate the homogeneity index among five MFM experts, and analyzed the results using Pearson correlation coefficients (Fig. 3A) and Spearman correlation coefficients (Fig. 3B). Overall, the results suggest that the raters exhibit a relatively high level of consensus when assessing the responses, showing a Homogeneity Index ranging from moderate to high. Specifically, the Pearson correlations ranged from 0.54 to 0.91, while the Spearman correlations ranged from 0.36 to 0.94.

**Figure 3.**
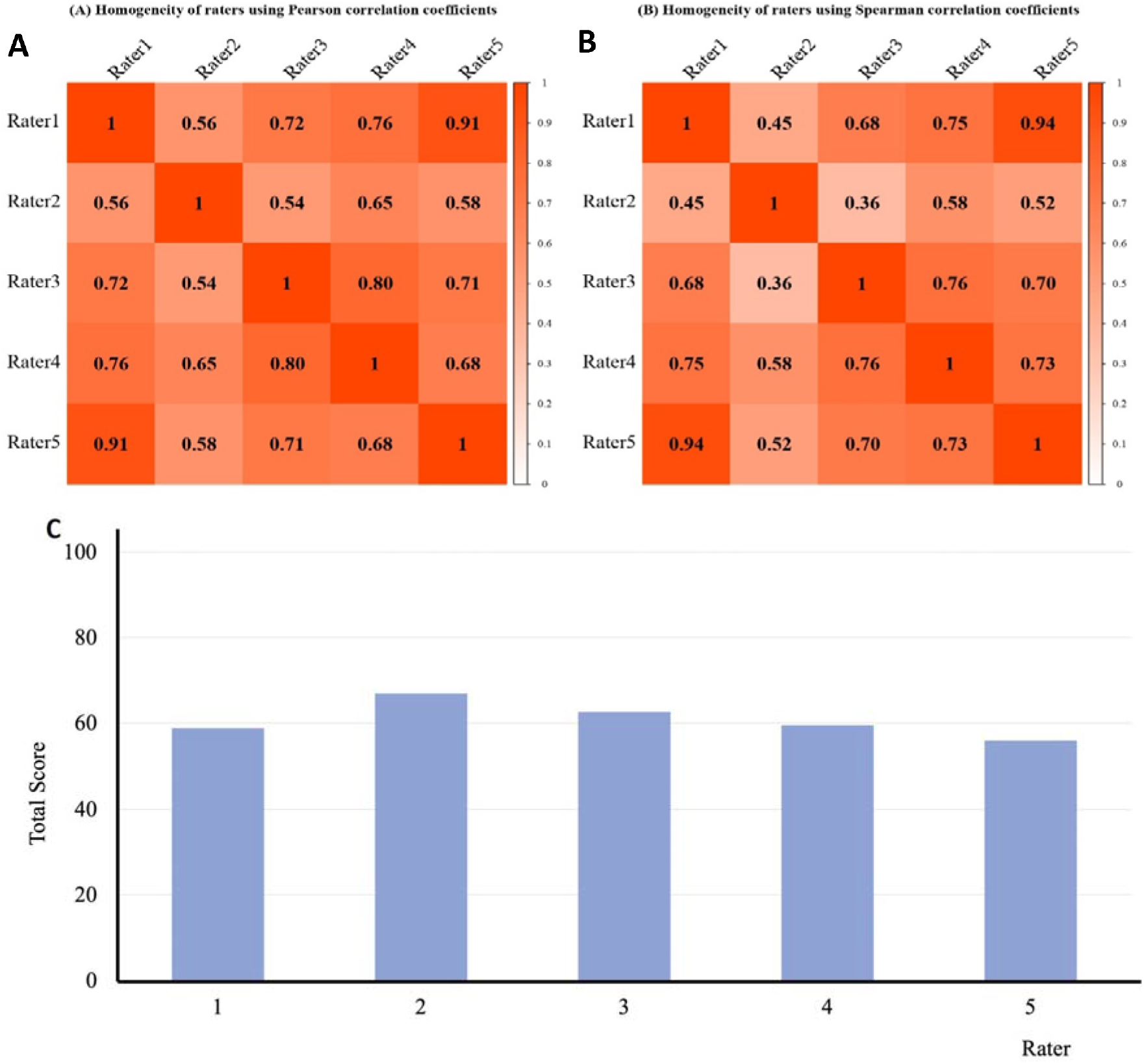
Scoring homogeneity among the raters. Using Pearson’s test (A) and Spearman’s test (B), we found medium-high homogeneity (0.54-0.91) & (0.36-0.94). The one-way ANOVA test with Scheffe’s post hoc analysis showed no difference among the 5 raters (C).

To further validate the reliability of the scoring process, a one-way ANOVA test with Scheffe’s post hoc analysis was used to examine the total scores for all responses (Fig. 3C). The result again showed that the variation among the raters was not significant (p> 0.05). Taken together, these results indicate that the raters had consistent views toward the CTG interpretations, indicating the evaluated results are dependable.

### The AI-LLMs showed variable performance in interpreting CTG images

AI-LLM, specifically CG4o, showed the best performance compared to other models, with a score of 77.86 (Fig. 4A). CG4o performed better than GemAdv and Copilot, which scored 57.14 and 47.29, respectively. These results indicate significant variations in performance among different AI-LLM systems in interpreting CTG data, with CG4o being the most competitive.

**Figure 4.**
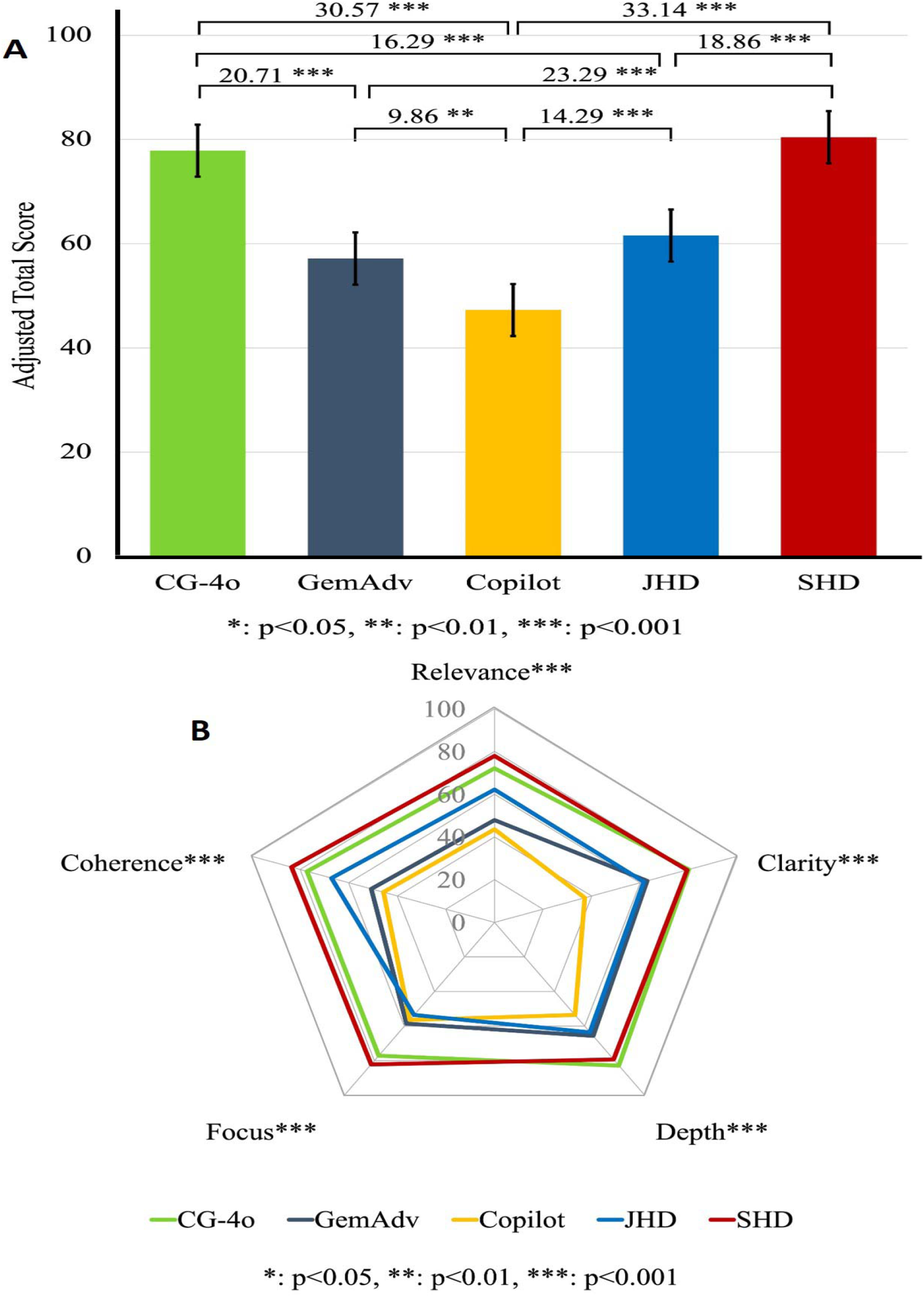
AI-LLMs and human doctors’ ability to interpret CTG. **(A)** CG4o scores significantly better than the other two AI-LLMs in providing CTG image interpretation. **(B)** The 5 parameters (relevance, clarity, depth, focus, and coherence) used for interpretation quality assessment show CG4o’s superiority significantly. JHD= Junior Human Doctor; SHD= Senior Human Doctor

In the evaluation of human doctors’ performance, senior doctors with over 10 years of experience achieved the highest score of 80.43, while junior doctors with less than 5 years of experience (JHD) scored 61.57. JHD scored lower than SHD and CG4o but still outperformed GemAdv and Copilot. Intriguingly, CG4o’s performance (77.86) only slightly trailed behind that of senior human doctors (80.43), showing no statistically significant difference, indicating the superior performance of CG4o in the interpretation of CTG.

A statistically significant difference (p < 0.001) was found between SHD and JHD, Copilot, and GemAdv, further indicating that senior doctors possess significantly superior expertise and experience in CTG analysis. Among the AI-LLM systems, CG4o also showed a statistically significant difference compared to GemAdv and Copilot (p < 0.001), reaffirming CG4o’s superiority among the AI-LLM models. JHD showed a significant difference (p < 0.001) compared to Copilot, this result suggests that human expertise, even at the junior level, still surpasses some AI models in complex clinical tasks.

The responses from the Chatbots and human doctors were further evaluated by six expert raters who used a 5-point Likert scale on relevance, clarity, depth, focus, and coherence (Fig. 4B). It is evident that SHD consistently ranked highest across all five parameters. This indicates that SHD provided more comprehensive CTG interpretations compared to AI-LLMs and JHD. JHD demonstrated competitive performance across most parameters. Although not reaching the same level as SHD, JHD consistently performed better than GemAdv and Copilot, particularly in coherence and clarity.

JHD demonstrated competitive performance across most parameters. Although not reaching the same level as SHD, JHD consistently performed better than GemAdv and Copilot, particularly in coherence and clarity. When comparing all AI-LLM models and human doctors, statistically significant differences (p < 0.001) were shown in this pentagram (Fig. 4B), suggesting a clear distinction between the tested groups. CG4o excelled in all assessment parameters compared to GemAdv and Copilot and even surpassed SHD in the depth parameter.

## Discussion

In this study, we assessed the abilities of three large language models in interpreting CTG images. This is the first report on using AI-LLM chatbots to interpret CTG results to the best of our knowledge. CTG is a crucial method for monitoring the fetus’s condition during pregnancy and labor. Its use helps in preventing complications that could be harmful to both the mother and the fetus. CTG can detect conditions such as hypoxia, fetal heart rate decelerations, and uterine rupture. Interpreting CTG requires the specialized expertise and experience of a physician.

The participation of two groups of doctors (SHD and JHD) allows us to examine how well AI-LLMs perform when compared to human doctors. The evaluation of the CTG image interpretations provided by the AI-LLMs and human doctors was conducted by five obstetrics experts. To ensure inter-rater consistency, we performed three statistical analyses (Pearson’s test, Spearman’s test, and the one-way ANOVA test with Scheffe’s post hoc analysis) to measure the homogeneity of the assessments (Fig. 3A-C). The studies showed a moderate to high homogeneity index among the raters, indicating a high agreement in their perception of the quality of CTG interpretations from the AI-LLMs. Although there were some variations in the ratings provided by the evaluators, these variances were not statistically significant (Fig. 3C), suggesting that their judgments were objective and consistent. Moreover, to ensure impartiality, the chatbot responses were coded, randomized, and then blindly assessed by the raters without knowing which chatbot generated each result. This further enhanced the confidence in the reliability of the AI-LLM evaluations, indicating that the assessments were not significantly influenced by individual biases among the raters.

We evaluated three AI-LLMs based on five key parameters: relevance, clarity, depth, focus, and coherence. The performance analysis revealed that CG4o outperformed GemAdv and Copilot in CTG image interpretation (Fig. 4A). The difference in scores between CG4o and GemAdv was 20.71, between CG4o and Copilot was 30.57, and between GemAdv and Copilot was 9.86. CG4o achieved the highest scores across all evaluation parameters (p<0.001) (Fig. 4B). These results indicate that among these three LMMs, CG4o provides the most consistent and accurate performance in CTG interpretation.

Intriguingly, comparisons of the CGT interpretations conducted by LLMs and human doctors indicate that CG4o show promising potential in assisting CTG interpretation. CG4o surpassed junior doctors and only slightly trailed behind senior doctors by 2.57, showing no statistical significance. These results show that CG4o holds great potential in interpreting CGT results. Nevertheless, the expert raters in this study agreed that senior doctors remain superior to LMMs in terms of relevance, coherence, and focus when providing CTG interpretations. This emphasizes the need for AI-LLM systems to be validated and supervised in their role as CTG image interpreters.

The lower performance of GemAdv and Copilot also highlights the need for further improvement in developing algorithms capable of achieving accuracy comparable to that of human doctors, particularly in highly complex contexts such as maternal health. On the other hand, CG4o can be used to assist less experienced doctors in interpreting CTG images. Improved diagnostic accuracy based on CTG results will ultimately lead to better patient care while also minimizing errors and complications in treatment. Despite CG4o’s performance, which is almost as good as that of experienced doctors (SHD) and even higher in “depth” parameters, there are numerous prospects for its use in patient care.

An advanced AI language model can be a valuable resource for medical personnel, especially in situations like fetal monitoring that require quick assessments. Integrating LLMs such as CG4o into clinical practice may reduce physicians’ workloads, speed up diagnostics, and enhance accuracy[16, 17]. These systems can improve patient care quality, reduce diagnostic errors, and enhance operational efficiency, making them valuable tools for medical professionals, especially in time-sensitive scenarios like fetal monitoring [16]. Leveraging AI-LLMs can enable healthcare providers to obtain quick and reliable assessments, improving patient outcomes. However, it is essential to address ethical concerns and adhere to safety guidelines in implementation[18, 19].

This study demonstrates that the AI-LLM system, particularly CG4o, can improve obstetric care by providing accurate, objective, and satisfactory interpretations. CG4o’s consistently high performance across key evaluation parameters suggests it can assist clinicians in making better-informed decisions, potentially reducing diagnostic errors, and improving patient outcomes. Since interpreting CTG requires the specialized expertise and experience of a physician, using LLMs is particularly valuable in settings where expert interpretation may not be readily available, ensuring a high standard of care across different healthcare environments.

However, while the potential benefits of integrating AI-LLMs into clinical practice are clear, careful consideration is needed regarding their ethical and operational implications. Ensuring patient privacy, and data security, and addressing algorithmic bias are crucial for the successful implementation of these systems. Moreover, training clinicians to use AI-LLM effectively and establishing clear guidelines for their use will be essential for these tools to complement, rather than replace, human expertise in healthcare.

In summary, CG4o demonstrated outstanding performance in interpreting CGT, making it an excellent tool to support human doctors in CTG interpretation. Its performance is only slightly inferior to that of SHD (p>0.05, indicating no statistical significance) and significantly outperforms other groups (p<0.05). However, it is important to emphasize that the role of the doctor as a validator and supervisor remains crucial, as the LLM does not operate autonomously and may provide imperfect answers.

## Limitations of the study

Despite the promising results, this study has several limitations. First, the evaluation was conducted using a relatively small sample size of CTG images, which may limit the generalizability of the findings, as AI-LLM may perform differently when exposed to more extensive and diverse CTG data. Another limitation lies in the potential bias caused by the selection of evaluation parameters. While the parameters of relevance, clarity, depth, focus, and coherence were carefully designed and tested in our previous AI-LMM studies[12, 15, 20]and are essential for assessing AI-LLM performance, they may not cover all aspects of effective CTG interpretation. Other important factors, such as the ability of AI-LLM to detect rare but clinically significant patterns or its adaptability to evolving clinical guidelines, were not explored in this study.

## Conclusion

This study evaluated the performance of three AI-LLMs—CG4o, GemAdv, and Copilot—in interpreting CTG images and compared them with senior (SHD) and junior doctors (JHD). The results showed that CG4o outperformed the other AI models, closely approaching the performance of senior doctors, particularly in the depth of analysis parameter. However, SHD still exhibited the best performance in terms of relevance, coherence, and focus, emphasizing that human expertise remains crucial. These findings suggest that AI-LLMs, especially CG4o, have the potential to assist doctors, particularly less experienced ones, in CTG interpretation. However, their use should be supervised by specialists to ensure patient safety and accuracy. This study highlights the benefits of AI-LLMs in enhancing obstetric care quality but also underscores the need for further research to ensure the reliability of these models in more complex clinical scenarios. In conclusion, the integration of advanced AI-LLMs, such as CG4o, into clinical practice aims to yield substantial benefits in the field of obstetrics, especially fetal monitoring in labor. Accurate and consistent application of AI-LLMs such as CG4o has the potential to improve clinical outcomes and operational efficiency thereby improving patient outcomes and avoiding morbidity and mortality.

## Data Availability

We have ensured that all important data required has been included in the Supplementary file. The exception is the raw values provided by individual doctors, which can be provided upon request.

## Authors’ contributions

KEG, LNL, and MT contributed to the conception of the study, methodology, and study design.

KEG contributed to data curation and validation.

LNL, YCH, and ZYY contributed to the statistical analysis of the data.

LNL, MT, and KEG contributed to visualization, which included figures, charts, and tables of data.

KEG contributed to project administration and resources.

KEG, MPW, MIAA, ASP, DPPB, RSM, and IF contributed to question testing and validation.

KEG, MPW, MIAA, ASP, DPPB, RSM, and IF contributed to question-and-answer scoring

KEG, CHL, LNL, and MT contributed to the manuscript writing and revising.

KEG and MT were responsible for the decision to submit the manuscript.

Supervision of this research, which includes responsibility for the research activity planning and execution, was overseen by MT.

All authors read and approved the final version of the manuscript.

## Acknowledgments

The authors are grateful for our collaborators, Ach S. Faridzi, MD (dr. Mohammad Zyn General Hospital, Sampang, Indonesia), Gallaran Matu, MD (dr. Soemarno Sosroatmodjo General Hospital, Bulungan, Indonesia), Nurlaella I. Nusi, MD (dr. Mohamad Soewandhie General Hospital, Surabaya, Indonesia), Roziana, MD (dr. Zainoel Abidin General Hospital, Banda Aceh, Indonesia), Yuni Setiawaty, MD (dr. Wahidin Sudiro Husodo General Hospital, Mojokerto, Indonesia). We also like to gratitude Bayu A.S. Putra, MD; Prita A. Malinda, MD; Ni Nyoman A.R. Pradnyani, MD; Rizqy Rahmatyah, MD; Royan M. Varendra, MD (Department of Obstetrics and Gynecology, Faculty of Medicine, Airlangga University, Surabaya, Indonesia).

## Declaration of generative AI and AI-assisted technologies in the writing process

During the preparation of this work, the authors used ChatGPT-4 and Grammarly to edit and proofread the manuscript to improve readability. After using these tools/services, the authors reviewed, verified, and edited the content as needed. The authors take full responsibility for the content of the publication.

## REFERENCES

1. Grivell, R. M.; Alfirevic, Z.; Gyte, G. M., and Devane, D., Antenatal cardiotocography for fetal assessment. Cochrane Database Syst Rev, 2015. 2015(9): p. Cd007863.

2. Caning, M. M.; Thisted, D. L. A.; Amer-Wählin, I.; Laier, G. H., and Krebs, L., Interobserver agreement in analysis of cardiotocograms recorded during trial of labor after cesarean. J Matern Fetal Neonatal Med, 2019. 32(22): p. 3778–83.

3. Cömert, Z.; Şengür, A.; Budak, Ü., and Kocamaz, A. F., Prediction of intrapartum fetal hypoxia considering feature selection algorithms and machine learning models. Health Inf Sci Syst, 2019. 7(1): p. 17.

4. Tarvonen, M.; Sainio, S.; Hämäläinen, E.; Hiilesmaa, V.; Andersson, S., and Teramo, K., Saltatory Pattern of Fetal Heart Rate during Labor Is a Sign of Fetal Hypoxia. Neonatology, 2020. 117(1): p. 111–7.

5. Ranaei-Zamani, N.; David, A. L.; Siassakos, D.; Dadhwal, V.; Aughwane, R.; Russell-Buckland, J.; Tachtsidis, I.; Hillman, S.; Melbourne, A., and Mitra, S., Saving babies and families from preventable harm: a review of the current state of fetoplacental monitoring and emerging opportunities. npj Women’s Health, 2024. 2(10).

6. Hartka, T., The American Journal of Emergency Medicine’s policy on large language model usage in manuscript preparation: Balancing innovation and responsibility. Am J Emerg Med, 2024. 82: p. 105–6.

7. Rios-Hoyo, A.; Shan, N. L.; Li, A.; Pearson, A. T.; Pusztai, L., and Howard, F. M., Evaluation of large language models as a diagnostic aid for complex medical cases. Front Med (Lausanne), 2024. 11: p. 1380148.

8. Gordon, E. B.; Towbin, A. J.; Wingrove, P.; Shafique, U.; Haas, B.; Kitts, A. B.; Feldman, J., and Furlan, A., Enhancing patient communication with Chat-GPT in radiology: evaluating the efficacy and readability of answers to common imaging-related questions. J Am Coll Radiol, 2023.

9. Rahsepar, A. A.; Tavakoli, N.; Kim, G. H. J.; Hassani, C.; Abtin, F., and Bedayat, A., How AI Responds to Common Lung Cancer Questions: ChatGPT vs Google Bard. Radiology, 2023. 307(5): p. e230922.

10. Wu, T.; He, S.; Liu, J.; Sun, S.; Liu, K.; Han, Q.-L., and Tang, Y., A Brief Overview of ChatGPT: The History, Status Quo and Potential Future Development. IEEE/CAA Journal of Automatica Sinica, 2023. 10(5): p. 1122–36.

11. Bhardwaz, S. and Kumar, J., An Extensive Comparative Analysis of Chatbot Technologies - ChatGPT, Google BARD and Microsoft Bing, in 2023 2nd International Conference on Applied Artificial Intelligence and Computing (ICAAIC). 2023. p. 673-9.

12. Gumilar, K. E.; Indraprasta, B. R.; Hsu, Y.-C.; Yu, Z.-Y.; Chen, H.; Irawan, B.; Tambunan, Z.; Wibowo, B. M.; Nugroho, H.; Tjokroprawiro, B. A., et al., Disparities in medical recommendations from AI-based chatbots across different countries/regions. Scientific Reports, 2024. 14(1).

13. Sikander, B.; Baker, J. J.; Deveci, C. D.; Lund, L., and Rosenberg, J., ChatGPT-4 and Human Researchers Are Equal in Writing Scientific Introduction Sections: A Blinded, Randomized, Non-inferiority Controlled Study. Cureus, 2023. 15(11): p. e49019.

14. Veras, M.; Dyer, J. O.; Rooney, M.; Barros Silva, P. G.; Rutherford, D., and Kairy, D., Usability and Efficacy of Artificial Intelligence Chatbots (ChatGPT) for Health Sciences Students: Protocol for a Crossover Randomized Controlled Trial. JMIR Res Protoc, 2023. 12: p. e51873.

15. Gumilar, K. E.; Indraprasta, B. R.; Faridzi, A. S.; Wibowo, B. M.; Herlambang, A.; Rahestyningtyas, E.; Irawan, B.; Tambunan, Z.; Bustomi, A. F.; Brahmantara, B. N., et al., Assessment of Large Language Models (LLMs) in Decision-Making Support for Gynecologic Oncology. Computational and Structural Biotechnology Journal, 2024.

16. Munley, C.; Jarmusch, A., and Chandrasekaran, S., LLM4VV: Developing LLM-driven testsuite for compiler validation. Future Generation Computer Systems, 2024. 160: p. 1–13.

17. Luo, M.-J.; Pang, J.; Bi, S.; Lai, Y.; Zhao, J.; Shang, Y.; Cui, T.; Yang, Y.; Lin, Z.; Zhao, L., et al., Development and Evaluation of a Retrieval-Augmented Large Language Model Framework for Ophthalmology. JAMA Ophthalmology, 2024.

18. Sblendorio, E.; Dentamaro, V.; Lo Cascio, A.; Germini, F.; Piredda, M., and Cicolini, G., Integrating human expertise & automated methods for a dynamic and multi-parametric evaluation of large language models’ feasibility in clinical decision-making. International Journal of Medical Informatics, 2024. 188: p. 105501.

19. Pashangpour, S. and Nejat, G., The Future of Intelligent Healthcare: A Systematic Analysis and Discussion on the Integration and Impact of Robots Using Large Language Models for Healthcare. Robotics, 2024. 13(8): p. 112.

20. Gumilar, K. E.; Ariani, G.; Wiratama, P. A.; Rimbun, R.; Yuliawati, T. H.; Chen, H.; Ibrahim, I. H.; Lin, C.-H.; Hung, T.-Y.; Anggrahini, D., et al., Assess the capabilities of AI-based large language models (AI-LLMs) in interpreting histopathological slides and scientific figures: performance evaluation study. JMIR Preprints, 2024.

